# Immature platelet dynamics are associated with clinical outcomes after major trauma

**DOI:** 10.1101/2023.08.25.23294628

**Authors:** Henry Schofield, Andrea Rossetto, Paul C Armstrong, Harriet E Allan, Timothy D Warner, Karim Brohi, Paul Vulliamy

**Author notes:** **Corresponding Author:** Dr Paul Vulliamy, Trauma Research Office, Ward 12D, Royal London Hospital, London, E1 1FR, United Kingdom. Henry Schofield and Andrea Rossetto contributed equally to this study. **Funding Information:** A.R. received a clinical research training fellowship (WT1390921) from the National Health Service Blood and Transplant (Bristol, UK). P.C.A., H.E.A. and T.D.W. received fundings from the British Heart Foundation. P.V. received fundings from the Academy of Medical Sciences and the National Institute for Health and Care Research. The Centre for Trauma Sciences receives support for rotational thromboelastometry reagents and equipment from Tem International GmbH (Munich, Germany). **Presentations:** Poster presentation at the XXX Congress of the International Society on Thrombosis and Hemostasis, London, UK, July 9-13, 2022.

## Abstract

**Background:** Major trauma results in dramatic changes in platelet behavior. Newly-formed platelets are more reactive than older platelets, but their contributions to hemostasis and thrombosis after severe injury have not been previously evaluated.

**Objectives:** To determine the relationship between immature platelet metrics and circulating drivers of platelet production with clinical outcomes after major injury.

**Methods:** Prospective observational cohort study of adult trauma patients. Platelet counts and the immature platelet fraction (IPF) were measured at admission, 24 hours, 72 hours and 7 days post-injury. Plasma proteins involved in thrombopoiesis were quantified at admission. The primary outcome was in-hospital mortality; secondary outcomes were venous thromboembolic events (VTE) and organ failure.

**Results:** At two hours post-injury, immature platelet counts (IPC) were significantly lower in non-survivors (n=40) compared to survivors (n=236; 7.3x10^9^/L vs 10.6x10^9^/L, p=0.009). Similarly, impaired platelet function on thromboelastometry was associated with a lower admission IPC (9.1x10^9^/L vs 11.9x10^9^/L, p<0.001). However, at later timepoints we observed a significantly higher IPF and IPC in patients who developed VTE (21.0x10^9^/L vs 11.1x10^9^/L, p=0.02) and prolonged organ dysfunction (20.9 x10^9^/L vs 11x10^9^/L, p=0.003) compared to those who did not develop complications. Plasma levels of thrombopoietin at admission were significantly lower in in non-survivors (p<0.001), in patients with organ failure (p<0.001) and in those who developed VTE (p=0.04).

**Conclusions:** Immature platelet depletion in the acute phase after major injury is associated with increased morality, whereas excessive immature platelet release at later timepoints may predispose to thrombosis and organ dysfunction.

**ESSENTIALS:**

- Newly-formed platelets are highly active, but their role in outcomes after major trauma is unknown
- Immature platelets were quantified in a cohort of 276 severely injured patients
- Early depletion of immature platelets is associated with increased mortality and coagulopathy
- Raised immature platelet at later timepoints increases the risk of thrombosis and organ failure

## Introduction

Platelet dysfunction is a central component of trauma-induced coagulopathy (TIC), a state of haemostatic incompetence which occurs in over 25% of severely injured patients and is associated with a four-fold increase in mortality[1,2]. A range of alterations in platelet form and function after major trauma have been described, including impaired responsiveness to *ex vivo* stimulation[3–5], loss of surface collagen receptors[6], and expansion of the procoagulant platelet subpopulation[7]. In patients who survive the bleeding phase, this initial coagulopathy switches towards a procoagulant state that predisposes to thrombotic events, which occur in up to a third of critically injured patients despite aggressive thromboprophylaxis[8]. Although our understanding of post-injury platelet biology has advanced significantly over the past decade, the precise mechanisms by which platelets contribute to TIC and their role in the subsequent prothrombotic switch are not fully understood.

Immature platelets are a highly reactive subpopulation that possess greater procoagulant and prothrombotic potential than older platelets[9]. These newly-formed platelets, also known as reticulated platelets, contain higher levels of messenger RNA which can be detected by nucleic acid dyes, enabling them to be distinguished from their older counterparts [10,11]. Commercially available assays for quantifying this immature platelet fraction (IPF) are widely available and have been primarily used to characterize thrombocytopenia, with a low IPF indicating a condition of decreased platelet production while a normal or increased IPF is suggestive of increased platelet destruction or consumption[12–14]. The IPF has clinical utility in predicting recovery from thrombocytopenia after chemotherapy[15], and more recently has been shown to provide prognostic information in patients with COVID-19 and sepsis, with higher IPF values being predictive of mortality[16,17]. However, immature platelets have not previously been studied in major trauma patients.

The objective of this study was to determine how immature platelet dynamics relate to clinical outcomes after major trauma. Given their procoagulant tendencies, we hypothesized that failure to mobilize immature platelets in the acute phase after injury would be associated with increased mortality due to bleeding, and that excessive production of immature platelets at later timepoints would increase the risk of thromboembolic events and organ failure. We also aimed to investigate the changes in circulating drivers of platelet production in the acute phase after major trauma.

## Methods

### Study design

We included patients recruited into the Activation of Coagulation and Inflammation in Trauma (ACIT)-II study at a single major trauma center. This is a platform prospective observational cohort study (REC reference 07/Q0603/29, ISRCTN12962642). Consent procedures and inclusion criteria have been described in detail previously[18]. For this analysis, patients meeting criteria for trauma team activation and had platelet count and IPF measurements at admission were eligible for inclusion. Patients who presented to hospital more than two hours after injury, were transferred from another hospital, had sustained burns over >5% of their body surface area, received more than two liters of fluid prior to recruitment, or who had a known bleeding diathesis were excluded.

### Study procedures

Blood samples were drawn on admission (within 2 hours of injury), at 24±2 hours, 72±12 hours and 7 days ±24 hours. Demographics, injury characteristics and outcomes were recorded prospectively by a trained member of the research team, and all patients were followed up on a daily basis until 28-days post-injury unless preceded by death or hospital discharge. The primary outcome of interest was in-hospital mortality. Secondary outcomes included 24-hour mortality, venous thromboembolism (VTE) and multiple organ dysfunction syndrome (MODS).

### Experimental methods

Platelet counts and the IPF were measured in our central hospital laboratory using a Sysmex XN-series analyzer (Sysmex, UK) according to standard operating procedures. From these variables we calculated the immature platelet count (IPC, as platelet count multiplied by IPF) and mature platelet count (MPC, as platelet count minus IPC) for each sample. Rotational thromboelastometry (RoTEM) was performed using a Delta instrument (TEM Innovations GmbH) within one hour of blood collection in accordance with the manufacturers’ instructions. Lactate and base deficit were measured with point-of-care blood gas analysis. For proteomic studies, blood was drawn into P100 vacutainers (Becton Dickinson, Plymouth, UK) and centrifuged at 2,500 *x* g for 10 minutes, the plasma fraction isolated and stored at - 80°C prior to analysis. Plasma proteins were measured using the SomaScan platform (SomaLogic, Inc., Boulder, CO) as previously described[19] and reported as relative fluorescence units (RFU).

### Definitions

Severe and critical injury were defined as an injury severity score (ISS) of >15 and >25 respectively. Previously established RoTEM cutoffs were used to define TIC (EXTEM CA5<40mm) and platelet dysfunction (EXTEM-FIBTEM CA5 <30mm)[20]. MODS was defined as a sequential organ failure assessment (SOFA) score of ≥6 on any day after the first day of admission and subdivided into early-resolving MODS (ERMODS) where the SOFA score fell to <6 within seven days of onset and prolonged MODS (PRMODS) when MODS persisted beyond seven days[21]. VTE was defined as either deep vein thrombosis or pulmonary embolism confirmed on imaging and subdivided into early (<7 days) and late (≥7 days) events. In some analyses we divided patients into groups according to the median platelet count (Count_LOW_ and Count_HIGH_) and median immature platelet fraction (IPF_LOW_ and IPF_HIGH_).

### Data analysis

Analysis was performed using Prism v8.0 (GraphPad Software Inc, San Diego, CA) and RStudio (R version 4.1.3 (2022-03-10)). Non-parametric data are reported as median with interquartile range (IQR) and were compared with Mann-Whitney U-test or Kruskal-Wallis test with Dunn’s post-test correction. Categorical data are reported as number and percentage and were compared with chi-squared or Fisher’s exact test, with correction for multiple comparisons using the Bonferroni method. Multivariable logistic regression models were constructed for main clinical outcomes (in-hospital mortality, all and late VTE events, MODS and PRMODS) and hemostatic parameters of coagulopathy and platelet dysfunction.

As confounding factors, we considered demographics variables (age and sex) and key descriptors of trauma severity (ISS and base deficit). No variables were considered as effect modifiers. All variables were included in the multivariable regressions independently of their statistical significance in the univariable analysis[22]. Due to the limited frequency of VTE events and PRMODS and to avoid the risk of model overfitting, we performed for these outcomes multiple multivariable regressions investigating the presence or absence of a consistent independent effect of IPC/IPF by adjusting for other confounding factors separately. Correlation between continuous variables was quantified with Spearman r. Survival times between groups were compared using log-rank test and presented as Kaplan-Meier curves. A two-tailed p-value of <0.05 was considered significant throughout and a complete case analysis was performed.

## Results

This study included 276 trauma patients recruited into the ACITII study between June 2020 and May 2022. The cohort comprised patients with a broad spectrum of injury severity (median ISS 13 [2-26]), of whom 86% (241/276) were male and 60% (166/276) had sustained a blunt mechanism of injury; detailed characteristics are presented in **supplemental table 1**. The median admission platelet count was 246x10^9^/L (199-299) and significant thrombocytopenia was rare (count <100 x10^9^/L: 14/276, 5%), in keeping with previous reports[3,23].

### Low immature platelet counts at admission are associated with increased mortality

We first examined the relationship between immature platelet metrics measured at admission with survival. Compared to survivors (n=236), non-survivors (n=40) had significantly lower immature platelet counts (7.3x10^9^/L [4.8-13.2] vs 10.6x10^9^/L [7.9-16.2], p=0.009), but the immature platelet fraction did not differ (4.0% [3.1-6.3] vs 4.7% [2.9-6.5], p=0.50). This pattern of immature platelet depletion in non-survivors persisted when we grouped patients according to injury severity, with markedly lower IPCs among non-survivors in patients with severe and critical injuries (**figure 1A)**. Similarly, we observed a lower IPC in patients who died within 24 hours of admission (5.0x10^9^/L [1.8=11.6] vs 10.4x10^9^/L [7.7-15.4], p=0.002; **supplemental figure 1A**), but again no differences in IPF (3.9% [2.5-4.4] vs 4.6% [2.9-6.5], p=0.14; **supplemental figure 1B**).

**Figure 1:**
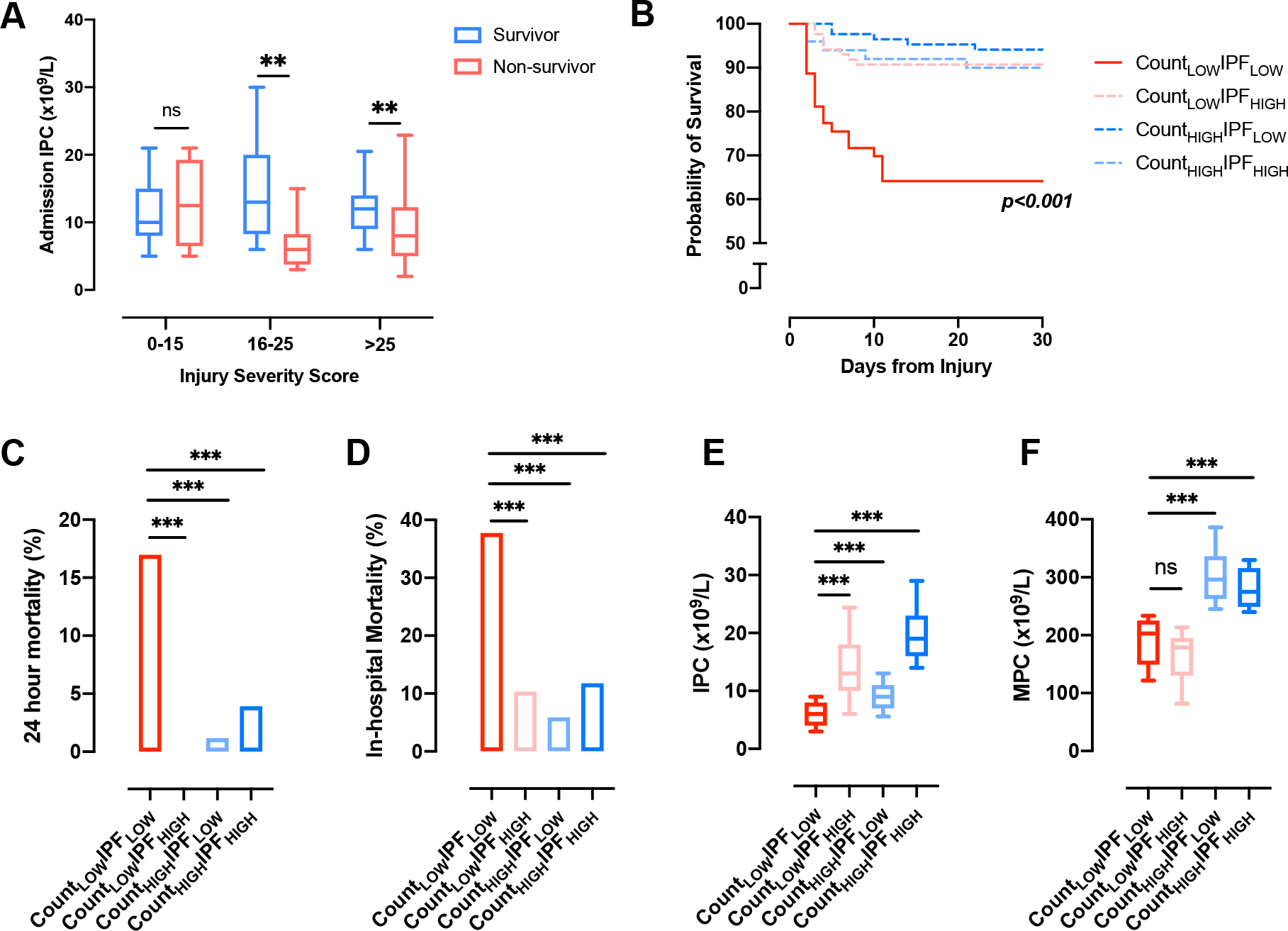
Admission immature platelet counts and mortality after major trauma. **A:** Immature platelet counts (IPC) in survivors and non-survivors with mild (injury severity score (ISS) 0-15), severe (ISS 16-25) and critical (ISS>25) injuries. **p<0.01, n.s., not significant, Mann-Whitney U-test. **B**: Kaplan-Meier curves for survival in patients grouped according to platelet count and immature platelet fraction (IPF). p-value derived using log-rank test. **C:** 24-hour mortality in patients grouped according to platelet count and IPF. **D**: In-hospital mortality. ***p<0.001, Fisher’s exact test. **E**: Immature platelet count. **F**: Mature platelet count (MPC). ***p<0.001, n.s., not significant, Kruskal-Wallis test with Dunn’s post-test correction. Box-whisker plots depict 10th-90th percentiles.

Consistent with this observation, when we stratified patients into two groups according to the median platelet count, we found that the mortality associated with lower platelet counts was concentrated in patients who also had a low IPF (Count_LOW_IPF_LOW_, n=53; **figure 1B**). Conversely, patients with lower platelet counts but a high IPF (Count_LOW_IPF_HIGH._ n=87) had a low mortality both at 24 hours (**figure 1C**) and overall (**figure 1D**), despite a similar incidence of critical injury in these two groups (34% vs 33%, p=0.99). Patients in the Count_LOW_IPF_LOW_ group had significantly lower immature platelet counts compared to the remainder of the cohort (**figure 1E**), but mature platelet counts were not different between Count_LOW_IPF_LOW_ and Count_LOW_IPF_HIGH_ (**figure 1F**). On multivariate regression analysis adjusting for key confounders (age, sex, ISS, base deficit), patients in the Count_LOW_IPF_LOW_ group had a 6.8-fold increase in odds of mortality (95% CI 2.4-19.2, p<0.001) compared to the remainder of the cohort (**table 1**). Collectively, these data suggest that depletion of circulating immature platelets is a key signature associated with increased mortality after major trauma, whereas maintained immature platelet counts appears to be protective.

**Table 1.** Multivariable regression analyses for in-hospital mortality.

|  | OR (95% CI) | p | Adj. OR (95% CI) | p |
| --- | --- | --- | --- | --- |
| Age, years | 1.025 (1.008-1.043) | 0.055 | 1.039 (1.013-1.065) | 0.003 |
| Sex, male | 1.177 (0.384-3.606) | 0.755 | 1.020 (0.260-3.994) | 0.978 |
| ISS | 1.094 (1.063-1.125) | <0.001 | 1.080 (1.043-1.119) | <0.001 |
| BD, mEq/L | 1.153 (1.090-1.220) | <0.001 | 1.084 (1.008-1.167) | 0.030 |
| COUNT<sub>LOW</sub>IPF<sub>LOW</sub> | 5.633 (2.592-12.24) | <0.001 | 6.801 (2.401-19.26) | <0.001 |

$R^2 = 0.51$ . OR, odds ratio; CI, confidence interval; Adj., adjusted; ISS, injury severity score; BD, base deficit; COUNT<sub>LOW</sub>IPF<sub>LOW</sub>, platelet count lower than the median and immature platelet count lower than the median.

### Patients with coagulopathy and platelet dysfunction have lower numbers of circulating immature platelets

Given that immature platelets have greater pro-aggregatory potential compared to mature platelets, we postulated that depletion of immature platelets could contribute to trauma-induced coagulopathy and platelet dysfunction. We compared patients with TIC (n=98) and without TIC (n=162) and found that coagulopathic patients had lower IPC levels (9.8x10^9^/L [5.6-12.7] vs 11.6x10^9^/L [8.4-18.5], p<0.001), a pattern which was also evident when we grouped patients according to injury severity (**figure 2A**) and when we stratified patients into Count_LOW_ and Count_HIGH_ groups as before (**figure 2B**). A similar picture was evident when we compared patients with (n=90) and without platelet dysfunction (n=166), with significantly lower IPC counts in those with platelet dysfunction (9.1x10^9^/L [5.5-12.6] vs 11.9x10^9^/L [8.7-18.5], p<0.001) which was again evident across injury subgroups (**figure 2C**) and platelet count subgroups (**figure 2D**). On multivariate regression, admission IPC was independently associated with TIC (p<0.001) and with platelet dysfunction (p<0.001) after adjusting for relevant confounders (**table 2)**.

**Table 2.** Multivariable logistic regression analysis of association between IPC and coagulopathy (top) and platelet dysfunction (bottom).

|  | OR (95% CI) | p | Adj. OR (95% CI) | p |
| --- | --- | --- | --- | --- |
| **EXTEM CA5 <40mm** |  |  |  |  |
| Age, years | 1.002 (0.988-1.016) | 0.786 | 0.999 (0.982-1.016) | 0.917 |
| Sex, male | 1.599 (0.691-3.702) | 0.184 | 1.956 (0.727-5.264) | 0.184 |
| ISS | 1.060 (1.037-1.083) | <0.001 | 1.051 (1.023-1.080) | <0.001 |
| BD, mEq/L | 1.163 (1.093-1.238) | <0.001 | 1.083 (1.010-1.161) | 0.026 |
| IPC, x10<sup>9</sup>/mL | 0.894 (0.848-0.944) | <0.001 | 0.897 (0.847-0.951) | <0.001 |
| **EXTEM-FIBTEM CA5 <30mm** |  |  |  |  |
| Age, years | 1.012 (0.998-1.026) | 0.106 | 1.011 (0.995-1.027) | 0.191 |
| Sex, male | 0.870 (0.392-1.930) | 0.732 | 0.946 (0.388-2.304) | 0.903 |
| ISS | 1.048 (1.027-1.070) | <0.001 | 1.032 (1.007-1.058) | 0.012 |
| BD, mEq/L | 1.124 (1.064-1.187) | <0.001 | 1.071 (1.007-1.139) | 0.030 |
| IPC, x10<sup>9</sup>/mL | 0.917 (0.872-0.963) | 0.001 | 0.926 (0.880-0.974) | 0.003 |

R<sup>2</sup> for EXTEM CA5 <40mm = 0.34. R<sup>2</sup> for EXTEM-FIBTEM CA5 <40mm = 0.24. EXTEM, tissue-factor activated rotational thromboelastometry; FIBTEM, tissue-factor activated rotational thromboelastometry with cytochalasin D; CA5, clot amplitude at 5 minutes; OR, odds ratio; CI, confidence interval; Adj., adjusted; ISS, injury severity score; BD, base deficit; IPC, immature platelet count.

**Figure 2:**
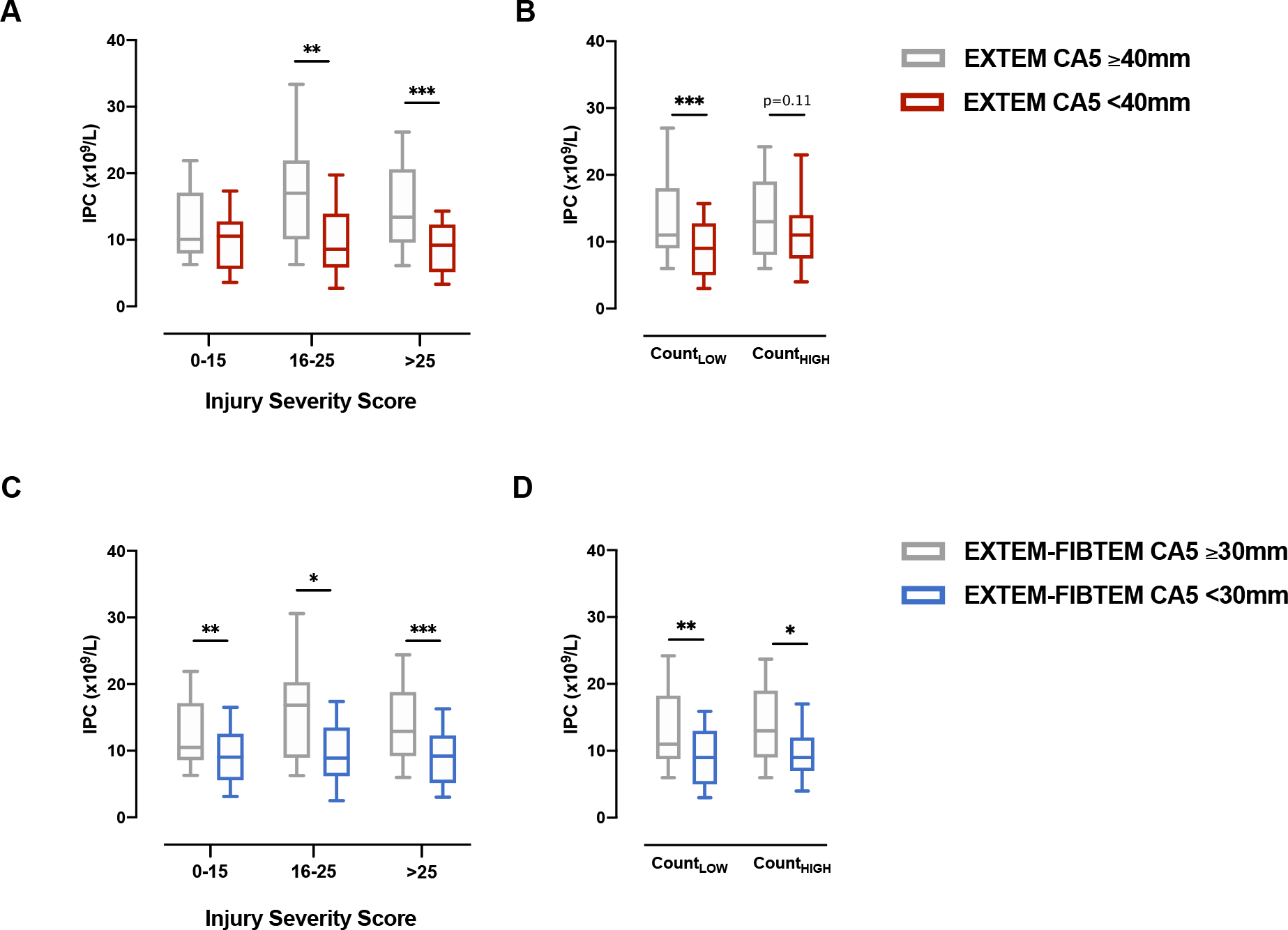
Immature platelet counts (IPC) in patients with coagulopathy and platelet dysfunction. **A**: IPC in patients with and without trauma-induced coagulopathy, defined as an EXTEM clot amplitude at 5 minutes (CA5) <40mm on rotational thromboelastometry (RoTEM), in patients with mild-moderate (injury severity score (ISS) 0-15), severe (ISS 16-25) and critical (ISS>25) injuries. **B**: IPC in patients stratified by the cohort median platelet count. **C:** IPC in patients with and without platelet dysfunction on RoTEM, defined as an EXTEM-FIBTEM CA5 <30mm, stratified by injury severity. **D**: IPC in patients with and without platelet dysfunction stratified by the cohort median platelet count. Box whisker plots depict 10-90^th^ percentiles. *p<0.05, **p<0.01, p<0.001, Mann-Whitney U-test.

### Increased immature platelets at later timepoints post-injury are associated with thrombosis and organ dysfunction

We next examined the relationship between immature platelet metrics and the risk of VTE and MODS. Patients who developed VTE (n=12) had a consistently higher IPF values over the first 7 days after injury (**supplemental figure 2A**) and a significantly higher peak IPF (10.3% [8.1-13.6] vs 5.0% [3.5-7.2], p<0.001) which was more pronounced in patients who developed VTE beyond the first week from injury (n=5; **figure 3A**). Peak IPCs were also significantly higher in patients who developed VTE (21.0x10^9^/L [10.3-32.6] vs 11.1x10^9^/L [8.0-18.0], p=0.02), particularly in those who developed late VTEs (**figure 3B**); this difference was primarily attributable to higher IPCs on day 7 rather than at earlier timepoints (**supplemental figure 2B**). The association between peak IPC with late VTE remained significant on regression analysis after adjusting for injury severity, shock and total platelet count (**supplemental table 2**).

**Figure 3:**
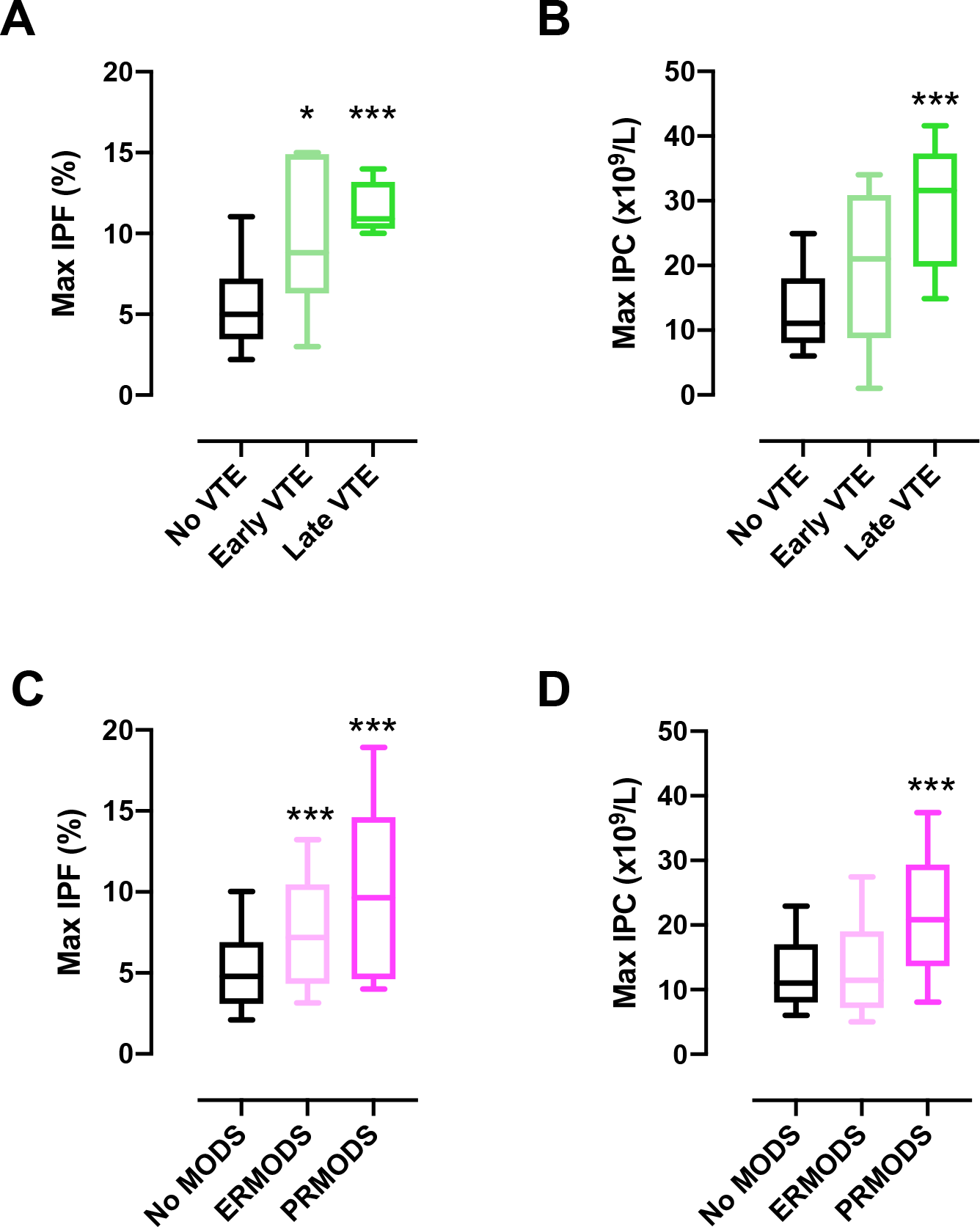
Immature platelet metrics in post-injury venous thromboembolism (VTE) and multiple organ dysfunction syndrome (MODS). **A and B**: Maximum Immature platelet fraction (IPF, A) and immature platelet count (IPC, B) over the first seven days post-injury in patients who did not develop VTE, developed VTE within the first week (early VTE) and developed VTE beyond the first week (late VTE). **C and D**: Maximum IPF (C) and IPC (D) over the first seven days post-injury in patients who did not develop MODS (n=), developed early resolving MODS (n=) and prolonged MODS (n=). Box-whisker plots depict 10-90^th^ percentiles. *p<0.05 ***p<0.001 vs no VTE/no MODS, Kruskal-Wallis test with Dunn’s post-test correction.

Similarly, patients who developed early-resolving MODS (ERMODS, n=52) and prolonged MODS (PRMODS, n=20) had significantly higher peak IPFs during the first 7 days after injury compared to those who did not develop MODS (**figure 3C** and **supplemental figure 3**). Maximum IPCs were higher in patients who developed PRMODS, but not in patients who developed ERMODS (**figure 3D**), which again was primarily attributable to increases in IPC at day 7 post-injury (**supplemental figure 3**). On regression analysis, peak IPC and IPF values were independently associated with PRMODS after adjusting for injury severity, shock and total/mature platelet count separately (**supplemental table 3)**.

### Rapid depletion of plasma thrombopoietin occurs in patients with severe injuries and adverse clinical outcomes

To investigate drivers of platelet production after major trauma, we analysed plasma proteomic data in blood samples drawn within two hours of injury from a separate cohort of patients recruited into the ACITII study (n=421; **supplemental table 4**). This cohort contained a similar spectrum of injury to our main cohort, with a median ISS of 11 (IQR 4-25) and a mortality of 5% (23/421). Plasma levels of thrombopoietin (TPO), the main driver of platelet production, were negatively correlated with injury severity score (*r*=-0.46, p<0.001), with the lowest levels in patients with critical injuries (**figure 4A**). A similar pattern was evident with plasma base deficit, a marker of systemic hypoperfusion, which showed a significant negative correlation with TPO (*r*=-0.29, p<0.001). Although TPO levels generally increase with a reduced platelet count due to lower availability of circulating TPO receptors[24], we found no correlation between TPO and platelet count (*r*=0.03, p=0.54; **figure 4B**), and indeed there was a non-significant trend towards lower TPO levels in patients with thrombocytopenia (n=35; 640 [544-783] vs 685 [593-779], p=0.12). In contrast, TPO levels were inversely proportional to plasma GPIbα, a marker of platelet consumption (*r*=-0.41, p<0.001; **figure 4C**). Plasma concentrations of the TPO receptor c-mpl showed no correlation with injury severity (*r*=0.10, p=0.95) and were not correlated with TPO levels (*r*=0.02, p=0.67, **supplemental figure 4**).

**Figure 4:**
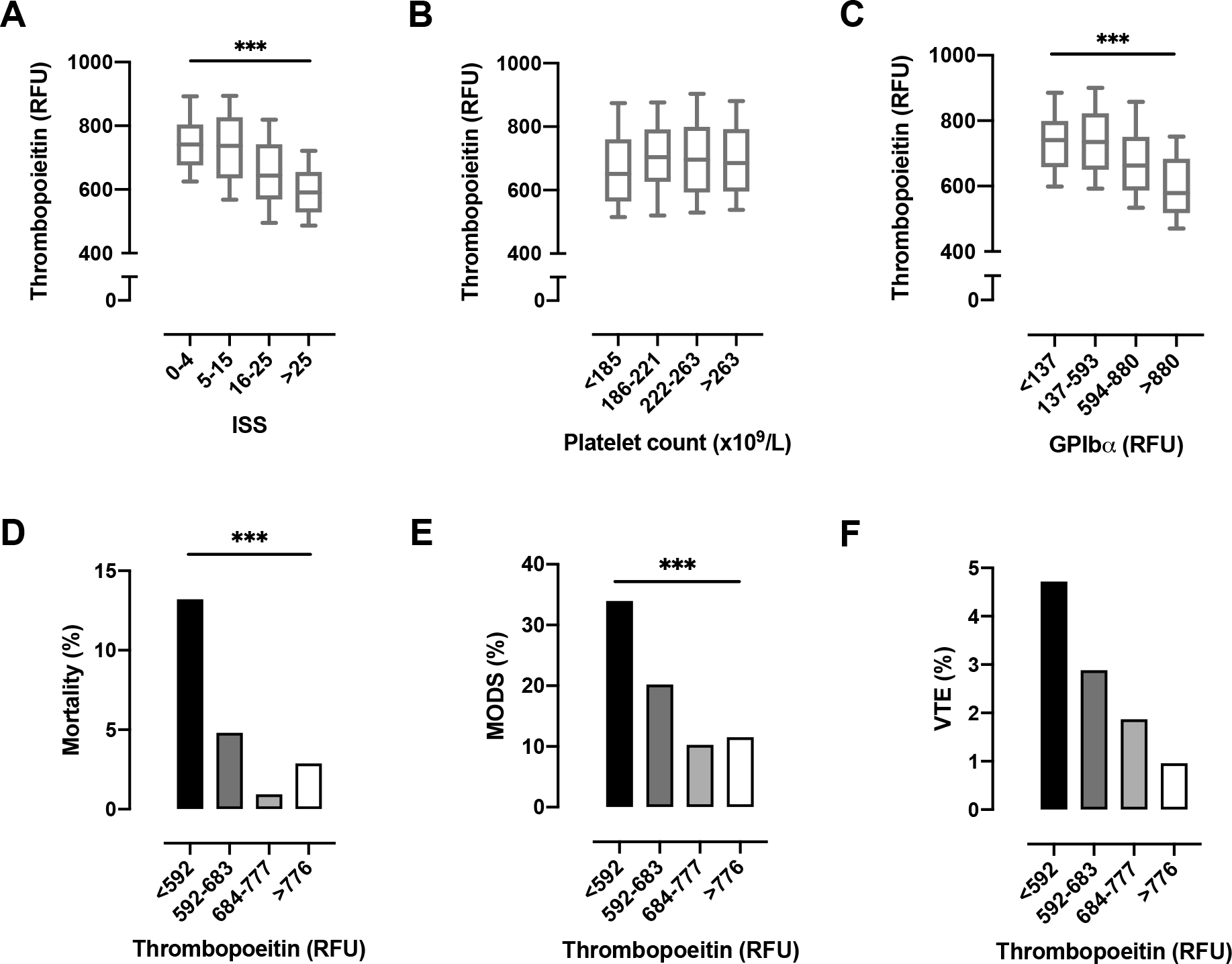
Admission plasma thrombopoietin (TPO) levels after major trauma. **A-C:** Levels of TPO in patients stratified by injury severity score (ISS, A), admission platelet count quartiles (B), and plasma GPIbα quartiles (C). RFU, relative fluorescence units. **D-E:** Mortality (D), multiple organ dysfunction syndrome (MODS, E) and venous thromboembolism (VTE, F) in patients stratified according to TPO quartiles. RFU, relative fluorescence units. ***p<0.001, Kruskal-Wallis test. Box whisker plots depict 10-90^th^ percentiles.

Finally, we examined the relationship between admission TPO levels with clinical outcomes. TPO concentrations were significantly lower in non-survivors compared to survivors (559 [483-659] vs 688 [595-5779], p<0.001), in patients with MODS compared to no MODS (616 [546-714] vs 737 [654-811], p<0.001) and those who developed VTE (628 (507-689 vs 683 [592-777], p=0.04). These differences were also evident when we stratified patients in quartiles according to the plasma TPO level, with the highest rates of mortality, MODS and VTE in the lowest quartile (**figure 4D-F**). These data indicate a mechanism of trauma-induced TPO depletion that occurs within hours of injury, is not attributable to increased TPO receptor levels in the circulation and is associated with poor outcomes.

## Discussion

In this cohort study of over 250 trauma patients, we describe important associations between the number and proportion of immature platelets with clinical outcomes. Our results demonstrate that early depletion of platelets together with a low immature platelet fraction is associated with a marked reduction in survival over the subsequent ten days. Conversely, a disproportionate increase in circulating immature platelets at later timepoints occurs in patients who develop organ dysfunction and thrombosis. Early depletion of plasma TPO levels occurs in proportion to injury severity and clinical outcome, which may indicate increased TPO uptake by platelet precursors in an attempt to compensate for tissue damage and blood loss. This represents a potential mechanism for subsequent increases in circulating immature platelets among patients who develop thrombotic and inflammatory complications.

Immature platelets are generally considered to have greater procoagulant and prothrombotic potential compared to aged platelets, acting as seeds for the assembly of platelet aggregates at sites of vessel injury[9,25]. Patients with major injuries may have multiple bleeding sites and frequently have an established coagulopathy that involves multiple aspects of hemostasis, including impairment of several key aspects of platelet haemostatic function[26]. Our results suggest that maintenance of a circulating pool of IPs in these patients may protect against coagulopathy and ongoing bleeding. Conversely, depletion of immature platelets below a certain threshold may contribute to compromised platelet haemostatic capacity, with a consequent increase in mortality.

The mechanism underlying this early immature platelet depletion in trauma may either be inadequate production by the bone marrow, increased consumption of immature platelets in developing thrombi, or a combination of both. The relative contributions of these two processes requires further study, but the fact that the mortality associated with IPC depletion is evident in patients who also have lower total platelet counts suggests that consumption is likely to be a contributor. However, our results show that depletion of non-immature platelets in isolation does not appear to confer an increased risk of mortality or coagulopathy, suggesting that immature platelets may help compensate for overall platelet depletion and coagulopathy until a certain threshold is reached.

Beyond the initial bleeding phase, trauma patients who survive rapidly shift from an initial coagulopathy towards a highly prothrombotic and proinflammatory state that contributes to adverse outcomes[27–29]. Our results suggest that whilst immature platelets may be protective during the bleeding phase, they may also drive development of subsequent complications in patients who survive to reach intensive care. Higher immature platelet counts are associated with an increased risk of thrombotic events in atherosclerosis[30,31], and diseases with increased platelet turnover such as diabetes mellitus carry an increased rate of thrombosis[32] Consistent with this, our results suggest that immature platelets may play a role in development of post-injury thrombosis. Platelets are increasingly recognized as important players in the pathophysiology of VTE[33], but are not specifically targeted by current thromboprophylactic regimens. Our findings add to the body of evidence implicating platelets in the development of post-traumatic VTE[33,34], and suggest that targeting platelets may enhance the efficacy of existing preventative measures. We also found that immature platelet numbers are elevated in patients who develop MODS, adding weight to existing evidence for a role for platelets beyond hemostasis and thrombosis in major trauma[35,36]. This is consistent with previous studies showing that a higher IPF/IPC is associated with worse outcomes in sepsis and COVID-19[16,17]. Further investigation into the characteristics of immature platelets released after injury, and the mechanisms by which they may contribute to post-injury organ dysfunction and thrombosis, could inform development of new prophylactic and therapeutic strategies.

The primary mediator of platelet production is TPO, which is produced by the liver and is essential for the survival and differentiation of megakaryocytes (MKs)[37]. We found that TPO levels were depleted within two hours of major trauma, with lower levels in patients with critical injuries and shock. Given that the half-life of TPO is 20-40 hours[38], this is unlikely to reflect synthetic failure. On binding to its receptor c-mpl, expressed on platelets and MKs, TPO is rapidly internalized and metabolized, providing a mechanism by which circulating levels of TPO fall[39]. We did not find evidence of increased c-mpl in circulation and TPO concentrations were not related to platelet count in trauma patients. Therefore, a potential explanation for the TPO depletion in plasma is increased uptake by MKs and their precursors via increased expression of surface c-mpl, although the mechanisms responsible for this are unclear. Given that TPO takes approximately 5 days to produce new platelets, this would explain the delayed rise in immature platelet counts that we observed in patients who subsequently developed thrombotic events and MODS.

An important limitation of this work is that while we describe quantitative changes in immature platelets, we have not explored the characteristics of newly formed platelets released after injury. A pertinent question for further study is whether major injury and blood loss induce a change in the properties of newly formed platelets released after injury. In addition, by virtue of the observational design of our study we are only able to describe associations rather than causal relationships, and further mechanistic work is needed to build on our findings reported here. Finally, we were not able to quantify molecular drivers of platelet production in the cohort in which immature platelet metrics were measured and therefore were not able to draw direct links between these variables.

In conclusion, this study provides new insights into immature platelet kinetics after major trauma and demonstrates their relevance to both early and late outcomes after injury. Based on the data reported here, we propose a model of initial immature platelet depletion in a subgroup of critically injured patients that is associated with mortality, followed by an excessive release of immature platelets at later timepoints that predisposes to thrombosis and organ dysfunction in those who survive the initial bleeding phase. These findings highlight the need for further work to define the mechanisms responsible for these changes and characterize the properties of newly-formed platelets released after major injury.

## Data Availability

All data produced in the present study are available upon reasonable request to the authors

## AUTHOR CONTRIBUTIONS

HS, AR, PV and KB designed the study. HS, AR, PV and PCA analysed the data and wrote the manuscript. HEA, PCA, TDW and KB critically revised the intellectual content. All authors have read and approved the final version of the manuscript.

## ACKOWLEDGMENTS

The authors thank Laura Green and Sean Platton for their support in liaising with the technicians at the central hospital laboratory.

