## Supplemental FIle 1 for "Immature platelet dynamics are associated with clinical outcomes after major trauma"

### SUPPLEMENTAL MATERIAL

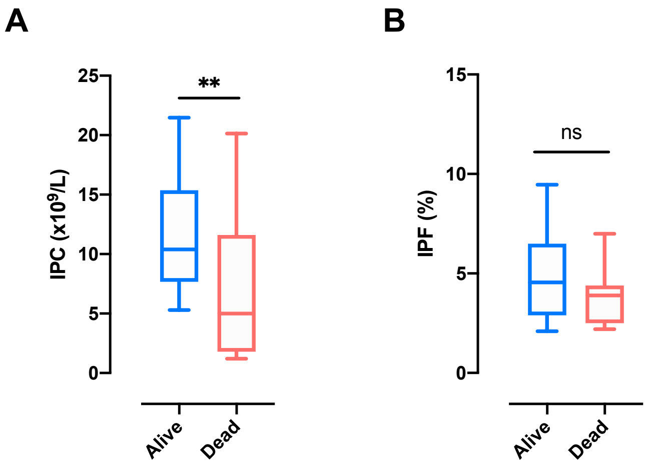


**Supplemental Figure 1: Immature platelet counts (A) and Immature platelet fraction (B) in survivors and non-survivors at 24 hours post-injury.** Box plots depict 10th-90th percentiles. **p<0.01, ns, not significant.

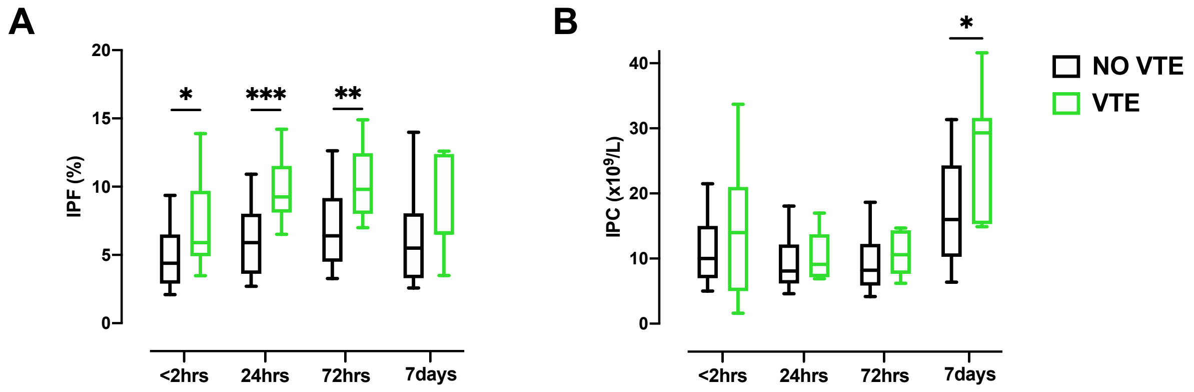


**Supplemental figure 2: Immature platelet metrics over time in patients with and without VTE.** A: immature platelet fraction. B: Immature platelet count. Box plots depict 10th-90th percentiles. *p<0.05,** p<0.01 ***p<0.001, Mann-Whitney U-test.

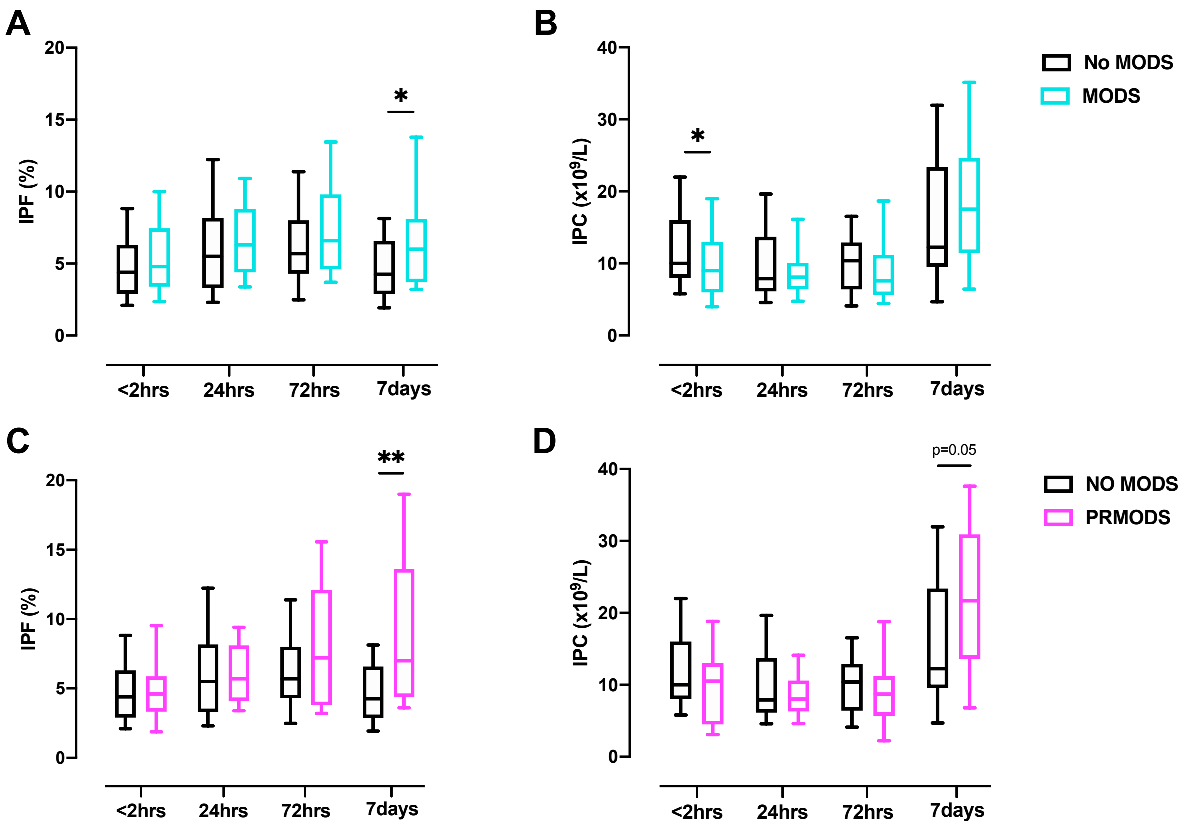


**Supplemental figure 3: Immature platelet metrics over time in patients with and without multiple organ dysfunction syndrome (MODS).** Box plots depict 10$^{th}$-90$^{th}$ percentiles. *p<0.05,** p<0.01, Mann-Whitney U-test.

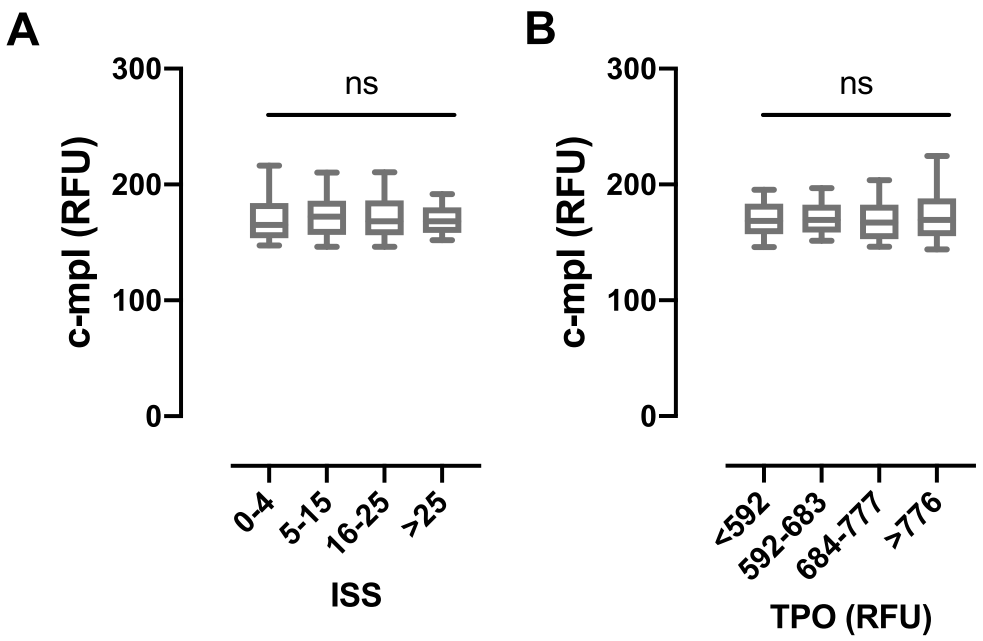


**Supplemental figure 4: Plasma levels of the thrombopoeitin receptor c-mpl in trauma patients. A**: c-mpl levels in patients stratified according to injury severity score (ISS). **B**: c-mpl levels in patients stratified in thrombopoeitin (TPO) quartiles. RFU, relative fluorescence units. ns, not significant. Box whisker plots depict median, interquartile range and 10-90$^{th}$ percentiles.

### Supplemental Table 1

**Supplemental Table 1. Characteristics of the study cohort.**

| | N = 276 |
|---|---|
| **Admission Demographics** | |
| Age (years) | 29 (18-44) |
| Female | 35 (14) |
| **Injury Description** | |
| Blunt mechanism | 166 (60) |
| ISS | 13 (2-26) |
| Head & Neck AIS ≥3 | 70 (30) |
| Thorax AIS ≥3 | 92 (39) |
| Abdomen & Pelvis AIS ≥3 | 50 (22) |
| Extremity AIS ≥3 | 54 (22) |
| **Admission Physiology** | |
| SBP <90 mmHg | 18 (8) |
| Heart rate >100 bpm | 120 (50) |
| Respiratory rate >22 bpm | 59 (27) |
| GCS <13 | 54 (28) |
| MHP activation | 100 (36) |
| **Admission Blood Results** | |
| IPF (%) | 4.6 (3.0-6.5) |
| IPC (x$10^9$/L) | 10 (7-15) |
| Platelet count (x$10^9$/L) | 246 (199-298) |
| Base deficit >6 mEq/L | 59 (23) |
| Lactate >6 mEq/L | 59 (23) |
| INR >1.2 | 35 (17) |
| EXTEM CA5 <40 mm | 97 (38) |
| **Outcomes** | |
| Mortality | 40 (14) |
| Length of stay | 5 (2-12) |
| RBC ≥4 in 24 hours | 34 (16) |
| MODS | 72 (28) |
| ERMODS | 52 (20) |
| PRMODS | 20 (8) |
| VTE | 12 (4) |

ISS, Injury severity score. AIS, abbreviated injury score. SBP, systolic blood pressure. GCS, Glasgow coma scale. IPF, immature platelet fraction. IPC, immature platelet count. INR, international normalised ratio. MODS, multiple organ dysfunction syndrome. ERMODS, early-resolving MODS. PRMODS, prolonged MODS. VTE, venous thromboembolism.

### Supplemental Table 2

**Supplemental Table 2. Characteristics of the plasma proteomics cohort.**

| | N = 421 |
|---|---|
| **Admission Demographics** | |
| Age (years) | 38 (28-53) |
| Female | 80 (19) |
| **Injury Description** | |
| Blunt mechanism | 415 (99) |
| ISS | 11 (4-25) |
| Head & Neck AIS ≥3 | 112 (27) |
| Thorax AIS ≥3 | 144 (34) |
| Abdomen & Pelvis AIS ≥3 | 37 (9) |
| Extremity AIS ≥3 | 112 (27) |
| **Admission Physiology** | |
| SBP <90 mmHg | 25 (6) |
| Heart rate >100 bpm | 131 (31) |
| Respiratory rate >22 bpm | 49 (14) |
| GCS <13 | 133 (32) |
| MHP activation | 48 (11) |
| **Admission Blood Results** | |
| Platelet count (x$10^9$/L) | 221 (185-262) |
| Base deficit >6 mEq/L | 39 (10) |
| Lactate >6 mEq/L | 26 (6) |
| INR >1.2 | 24 (7) |
| EXTEM CA5 <40 mm | 112 (28) |
| **Outcomes** | |
| Mortality | 23 (5) |
| RBC ≥4 in 24 hours | 38 (9) |
| MODS | 74 (18) |
| ERMODS | 53 (13) |
| PRMODS | 21 (5) |
| VTE | 11 (3) |

ISS, Injury severity score. AIS, abbreviated injury score. SBP, systolic blood pressure. GCS, Glasgow coma scale. INR, international normalised ratio. MODS, multiple organ dysfunction syndrome. ERMODS, early-resolving MODS. PRMODS, prolonged MODS. VTE, venous thromboembolism.

### Supplemental table 3

**Supplemental Table 3. Multivariable logistic regression analysis of association between IPC (top) and IPF (bottom) with incidence of all VTE and late VTE (developed later than 7 days) events.**

| | All VTE | | | | Late VTE | | | |
|---|---|---|---|---|---|---|---|---|
| | OR (95% CI) | p | Adj. OR (95% CI) | p | OR (95% CI) | p | Adj. OR (95% CI) | p |
| **Immature platelet count** | | | | | | | | |
| ISS | 1.028 (0.994-1.064) | 0.110 | 1.017 (0.980-1.056) | 0.381 | 1.035 (0.985-1.089) | 0.175 | 1.020 (0.961.1.082) | 0.517 |
| Max IPC, $x10^9/L$ | 1.094 (1.038-1.153) | 0.001 | 1.088 (1.031-1.148) | 0.002 | 1.136 (1.051-1.227) | 0.001 | 1.129 (1.044.1.222) | 0.002 |
| BD, mEq/L | 1.098 (1.026-1.175) | 0.007 | 1.112 (1.030-1.200) | 0.006 | 1.079 (0.978-1.191) | 0.131 | 1.092 (0.969-1.230) | 0.148 |
| Max IPC, $x10^9/L$ | 1.094 (1.038-1.153) | 0.001 | 1.089 (1.032-1.150) | 0.002 | 1.136 (1.051-1.227) | 0.001 | 1.133 (1.050-1.223) | 0.001 |
| Max MPC, $x10^9/L$ | 1.003 (0.999-1.007) | 0.150 | 1.001 (0.997-1.006) | 0.535 | 1.008 (1.003-1.012) | 0.001 | 1.006 (1.001-1.011) | 0.013 |
| Max IPC, $x10^9/L$ | 1.094 (1.038-1.153) | 0.001 | 1.090 (1.033-1.150) | 0.002 | 1.136 (1.051-1.227) | 0.001 | 1.125 (1.032-1.226) | 0.007 |
| **Immature platelet fraction** | | | | | | | | |
| ISS | 1.028 (0.994-1.064) | 0.110 | 1.013 (0.976-1.053) | 0.491 | 1.035 (0.985-1.089) | 0.175 | 1.018 (0.962-1.078) | 0.530 |
| Max IPF, % | 1.143 (1.043-1.253) | 0.004 | 1.127 (1.020-1.246) | 0.019 | 1.166 (1.027-1.324) | 0.018 | 1.145 (0.997-1.316) | 0.056 |
| BD, mEq/L | 1.098 (1.026-1.175) | 0.007 | 1.076 (0.998-1.161) | 0.056 | 1.079 (0.978-1.191) | 0.131 | 1.035 (0.924-1.160) | 0.555 |
| Max IPF, % | 1.143 (1.043-1.253) | 0.004 | 1.089 (0.980-1.209) | 0.112 | 1.166 (1.027-1.324) | 0.018 | 1.142 (0.990-1.317) | 0.069 |
| Max PLT, $x10^9/L$ | 1.003 (0.999-1.007) | 0.092 | 1.004 (1.001-1.008) | 0.026 | 1.008 (1.003-1.012) | 0.001 | 1.010 (1.004-1.015) | 0.001 |
| Max IPF, % | 1.143 (1.043-1.253) | 0.004 | 1.176 (1.064-1.299) | 0.001 | 1.166 (1.027-1.324) | 0.018 | 1.289 (1.078-1.542) | 0.005 |

IPC, immature platelet count; IPF, immature platelet fraction; VTE, venous thromboembolism; OR, odds ratio; CI, confidence interval; Adj., adjusted; ISS, injury severity score; BD, base deficit; MPC, mature platelet count; PLT, maximum platelet count.

### Supplemental table 4

**Supplemental Table 4. Multivariable logistic regression analysis of association between IPC (top) and IPF (bottom) with the development of MODS and PRMODS.**

| | MODS | | | | PRMODS | | | |
|---|---|---|---|---|---|---|---|---|
| | OR (95% CI) | p | Adj. OR (95% CI) | p | OR (95% CI) | p | Adj. OR (95% CI) | p |
| **Immature platelet count** | | | | | | | | |
| ISS | 1.121 (1.090-1.153) | <0.001 | 1.119 (1.088-1.151) | <0.001 | 1.084 (1.050-1.119) | <0.001 | 1.086 (1.048-1.125) | <0.001 |
| Max IPC, x10$^9$/L | 1.039 (1.007-1.072) | 0.018 | 1.018 (0.977-1.060) | 0.393 | 1.093 (1.044-1.144) | <0.001 | 1.090 (1.035-1.147) | 0.001 |
| BD, mEq/L | 1.176 (1.110-1.246) | <0.001 | 1.180 (1.113-1.250) | <0.001 | 1.092 (1.031-1.157) | 0.003 | 1.102 (1.034-1.174) | 0.003 |
| Max IPC, x10$^9$/L | 1.039 (1.007-1.072) | 0.018 | 1.048 (1.010-1.088) | 0.012 | 1.093 (1.044-1.144) | <0.001 | 1.081 (1.031-1.134) | 0.001 |
| Max MPC, x10$^9$/L | 0.997 (0.994-1.000) | 0.054 | 0.995 (0.992-0.999) | 0.008 | 1.000 (0.996-1.004) | 0.986 | 0.997 (0.992-1.002) | 0.292 |
| Max IPC, x10$^9$/L | 1.039 (1.007-1.072) | 0.018 | 1.055 (1.019-1.093) | 0.002 | 1.093 (1.044-1.144) | <0.001 | 1.102 (1.050-1.157) | <0.001 |
| **Immature platelet fraction** | | | | | | | | |
| ISS | 1.121 (1.090-1.153) | <0.001 | 1.114 (1.083-1.146) | <0.001 | 1.084 (1.050-1.119) | <0.001 | 1.077 (1.041-1.115) | <0.001 |
| Max IPF, % | 1.196 (1.113-1.285) | <0.001 | 1.139 (1.042-1.245) | 0.004 | 1.178 (1.084-1.128) | <0.001 | 1.135 (1.031-1.249) | 0.010 |
| BD, mEq/L | 1.176 (1.110-1.246) | <0.001 | 1.166 (1.099-1.238) | <0.001 | 1.092 (1.031-1.157) | 0.003 | 1.069 (1.001-1.140) | 0.045 |
| Max IPF, % | 1.196 (1.113-1.285) | <0.001 | 1.186 (1.091-1.289) | <0.001 | 1.178 (1.084-1.128) | <0.001 | 1.116 (1.017-1.226) | 0.021 |
| Max PLT, x10$^9$/L | 0.997 (0.995-1.000) | 0.091 | 0.999 (0.996-1.002) | 0.380 | 1.001 (0.997-1.005) | 0.753 | 1.002 (0.999-1.006) | 0.212 |
| Max IPF, % | 1.196 (1.113-1.285) | <0.001 | 1.192 (1.109-1.282) | <0.001 | 1.178 (1.084-1.128) | <0.001 | 1.195 (1.096-1.302) | <0.001 |

IPC, immature platelet count; IPF, immature platelet fraction; MODS, multiple organ dysfunction syndrome; PRMODS, prolonged multiple organ dysfunction syndrome; OR, odds ratio; CI, confidence interval; Adj., adjusted; ISS, injury severity score; BD, base deficit; MPC, mature platelet count; PLT, platelet count.
